# CTKG: A Knowledge Graph for Clinical Trials

**DOI:** 10.1101/2021.11.04.21265952

**Authors:** Ziqi Chen, Bo Peng, Vassilis N. Ioannidis, Mufei Li, George Karypis, Xia Ning

**Affiliations:** The Ohio State University; Amazon Web Services AI; Amazon Web Services Shanghai AI Lab

## Abstract

Effective and successful clinical trials are essential in developing new drugs and advancing new treatments. However, clinical trials are very expensive and easy to fail. The high cost and low success rate of clinical trials motivate research on inferring knowledge from existing clinical trials in innovative ways for designing future clinical trials. In this manuscript, we present our efforts on constructing the first publicly available Clinical Trials Knowledge Graph, denoted as `CTKG`. `CTKG` includes nodes representing medical entities in clinical trials (e.g., studies, drugs and conditions), and edges representing the relations among these entities (e.g., drugs used in studies). Our embedding analysis demonstrates the potential utilities of `CTKG` in various applications such as drug repurposing and similarity search, among others.

## Introduction

Clinical trials are studies aiming at determining the safety and efficacy of interventions, treatments or investigational drugs on human subjects^1^. Effective and successful clinical trials are essential in developing new drugs and advancing new treatments^2^. However, clinical trials are very expensive. As reported in Sertkaya *et al*.^3^, the average cost of a single phase in clinical trials ranges from 1.4 million up to 52.9 million US dollars. In addition, the success rate of the clinical trials is considerably low. As reported in Wong *et al*.^4^, for certain therapeutic groups like Oncology, the overall success rate of clinical trials could be as low as 3.4%. The high cost and low success rate of clinical trials motivate deliberate analysis of existing clinical trials, inferring knowledge from them, utilizing existing clinical trials in innovative ways, and accordingly carefully designing future clinical trials. The Access to Aggregate Content of ClinicalTrials.gov (AACT) database^5^ represents an effort in enhancing the accessibility and analysis of the clinical trial data. However, as a relational database, AACT is not formatted for the purpose of inferring new knowledge from existing clinical trials^6^. A Knowledge Graph (KG), instead, is a graph representation in which information entities are represented as nodes, and their relations are coded as edges connecting the corresponding nodes. In contrast to relational databases, KG has been proven^7–10^ to be an effective representation for knowledge inference purposes. Constructing a KG over clinical trial data is vital for advancing the analysis and research of clinical trials. In this manuscript, we present our work on constructing a such KG, referred to as Clinical Trials Knowledge Graph, denoted as `CTKG`, and also release `CTKG` to the research community to facilitate advanced research using clinical trial data. `CTKG` includes nodes representing medical entities (e.g., studies, drugs and conditions), and edges representing relations among these entities (e.g., drugs used in studies). Different from the recently released knowledge base^11^ that focuses only on extracting medical entities from the eligibility criteria in clinical trials, `CTKG` includes more medical entities (e.g., adverse events and outcomes) and also the relations among these entities. The rich information in `CTKG` could enable more biomedical applications (e.g., adverse drug event prediction, outcome prediction) than the existing knowledge base in clinical trials. Figure 1 presents the schema of `CTKG`. The detailed descriptions of nodes and edges in `CTKG` will be presented in Section “Nodes in `CTKG`”. To the best of our knowledge, `CTKG` is the first publicly available clinical trials knowledge graph in the scientific research community. The results of the embedding analysis over `CTKG` demonstrate its potential utilities in various applications such as drug repurposing and similarity search, among others.

**Figure 1.**
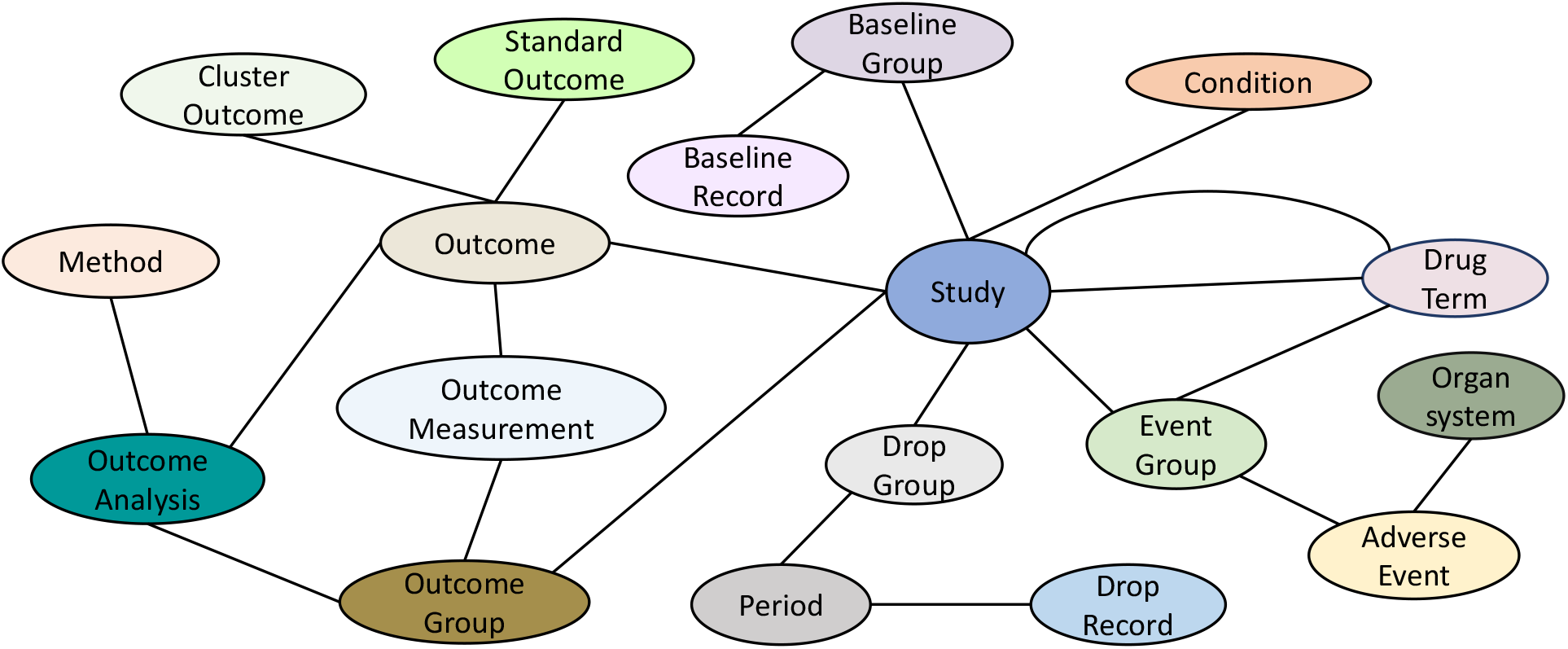
Schema of `CTKG`

## Results

### Clinical Trials Data

The clinical trials data in `CTKG` is collected from the Access to Aggregate Content of ClinicalTrials.gov (AACT) database. AACT is a publicly available relational database, which contains the information of every clinical trial registered in ClinicalTrials.gov, and is updated on a daily basis. In AACT, each clinical trial, also referred to as a study, is associated with a unique National Clinical Trial (NCT) ID, and all the information of a clinical trial is stored in 45 different tables. For example, information representing the medications, procedures and other actions provided or conducted in a clinical trial is stored in two tables: “interventions” and “browse interventions”; information representing the measurements used to evaluate the safety and efficacy of drugs or procedures studied in clinical trials is stored in the table “outcomes.” All the tables and their schemas are available in https://aact.ctti-clinicaltrials.org/. Currently, `CTKG` includes 8,210 studies registered in the ClinicalTrials.gov between 1999 and 2020, and will be updated with new studies in the future. It includes only studies that have drug interventions and reported outcomes.

Note that `CTKG` does not include all the tables in AACT. For example, `CTKG` does not include tables such as “Sponsors”, “Overall officials” and “Result contacts” because they are not directly related to the design and results of clinical trials, and including them may not significantly benefit the knowledge graph in analyzing the relations among medical entities. Other AACT tables such as “Provided documents” and “Documents” contain the links to detailed study protocols, informed consent forms and statistical analysis plans, etc. These documentations have rich textual information that might be complementary to the structural relations represented by `CTKG`. However, such information is highly specific to each individual clinical trial, and does not help establish new relations across clinical trials if no natural language processing is applied first, which by itself is highly non-trivial. Therefore, `CTKG` does not include such tables; instead, `CTKG` uses AACT’s original study IDs so that all such information can still be retrieved from AACT if needed. `CTKG` does not include other AACT tables such as “Calculated values”, “Design outcomes” and “Design group interventions” because information in such tables is already included in other tables that `CTKG` includes. Table 1 summarizes the AACT tables that are included and are not included in `CTKG`.

**Table 1.**
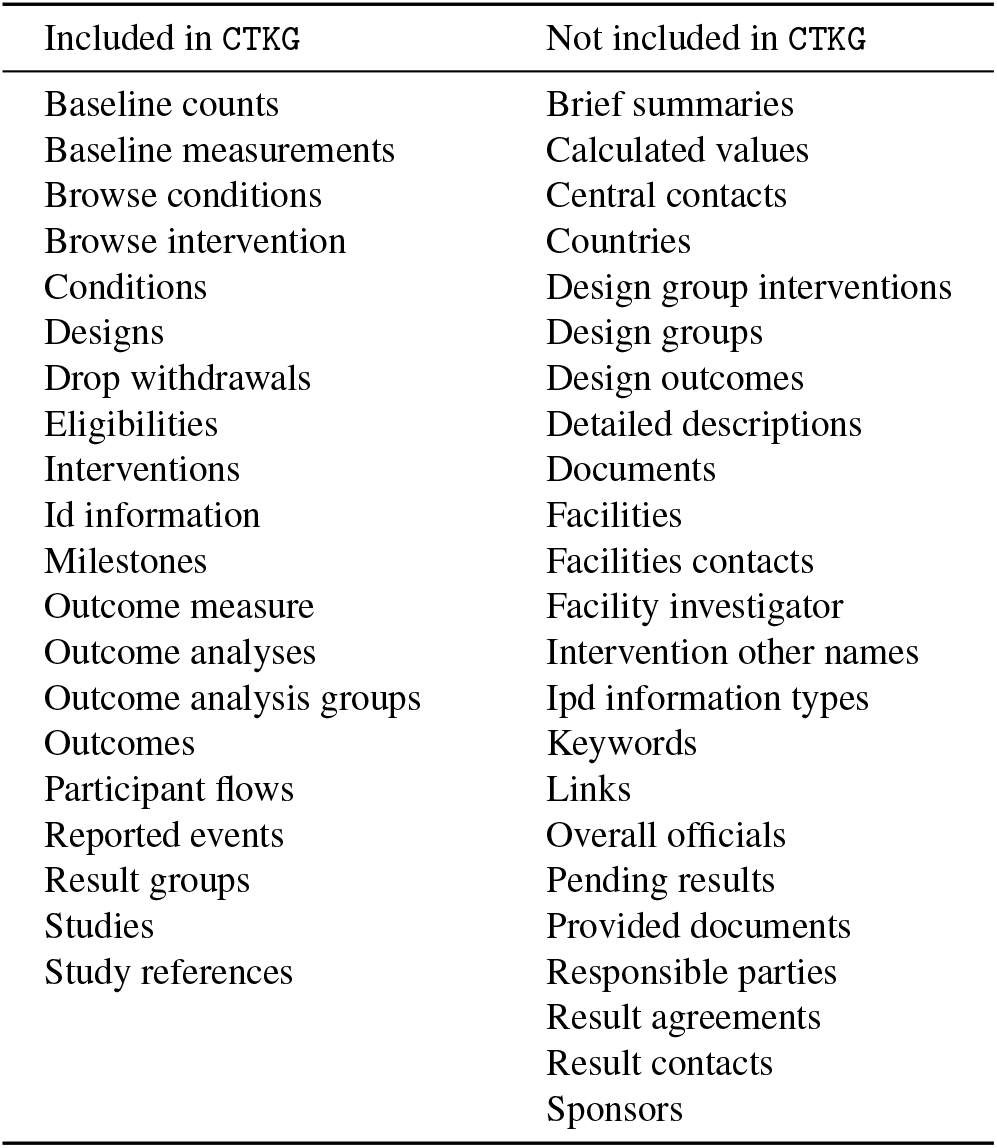
AACT tables included and not included in `CTKG`

### CTKG Schema

Figure 1 presents the schema of `CTKG`. The schema presents the different information entities involved in clinical trials, represented as nodes, and the relations among them, represented as edges. There are 18 types of nodes and 21 types of edges in `CTKG`. Each node and edge type has attributes describing the properties of the nodes and edges. The statistics of different nodes and edges are presented in Table 2 and Table 3, respectively. Detailed descriptions of node and edge attributes are available in the online documentation of `CTKG`: https://github.com/ninglab/CTKG/blob/main/Schema.pdf.

**Table 2.**
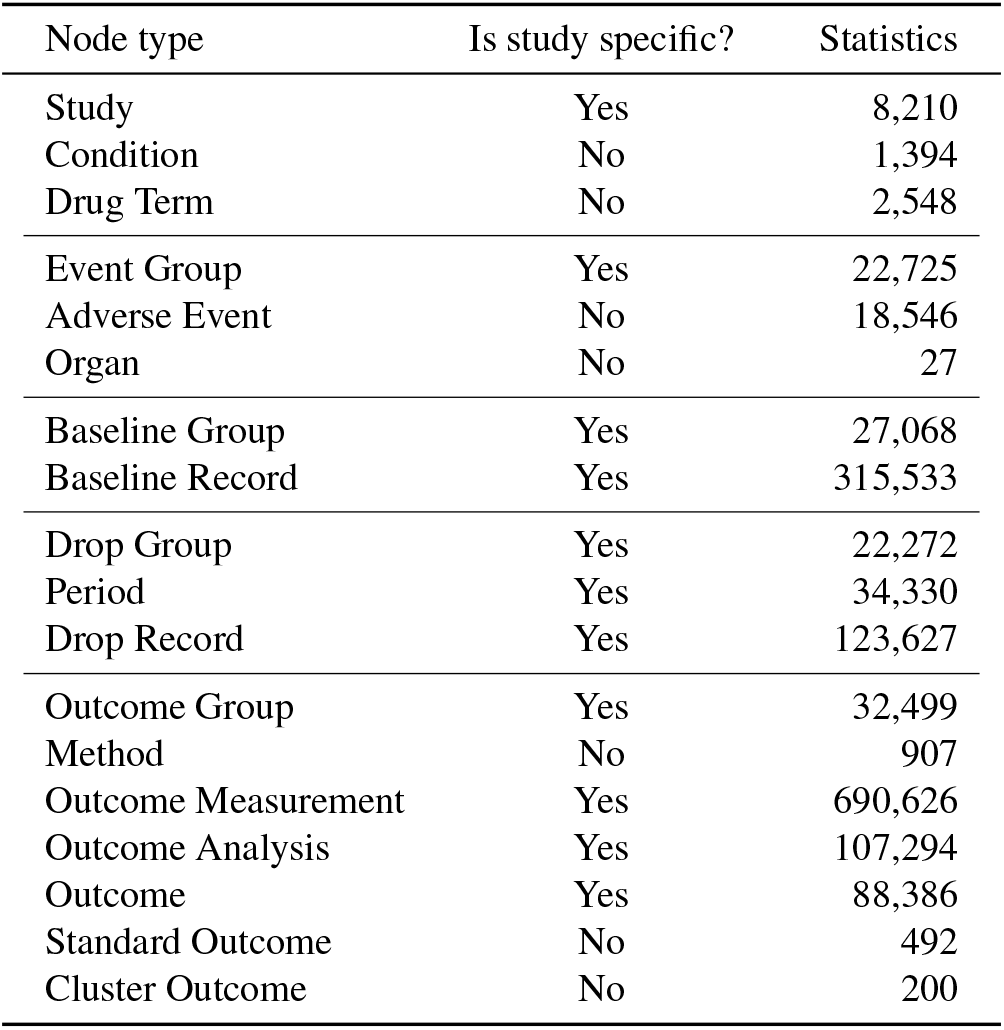
Statistics of node types in `CTKG`

| Node type | Is study specific? | Statistics |
| --- | --- | --- |
| Study | Yes | 8,210 |
| Condition | No | 1,394 |
| Drug Term | No | 2,548 |
| Event Group | Yes | 22,725 |
| Adverse Event | No | 18,546 |
| Organ | No | 27 |
| Baseline Group | Yes | 27,068 |
| Baseline Record | Yes | 315,533 |
| Drop Group | Yes | 22,272 |
| Period | Yes | 34,330 |
| Drop Record | Yes | 123,627 |
| Outcome Group | Yes | 32,499 |
| Method | No | 907 |
| Outcome Measurement | Yes | 690,626 |
| Outcome Analysis | Yes | 107,294 |
| Outcome | Yes | 88,386 |
| Standard Outcome | No | 492 |
| Cluster Outcome | No | 200 |

**Table 3.** Statistics of relation types in `CTKG`

| Relation type | Node type 1 | #Node 1 | Node type 2 | #Node 2 | #Relations |
| --- | --- | --- | --- | --- | --- |
| Study-Condition | Study | 8,210 | Condition | 1,394 | 17,259 |
| Study-EventGroup | Study | 8,172 | Event Group | 22,725 | 22,725 |
| Study-BaselineGroup | Study | 8,209 | Baseline Group | 27,068 | 27,068 |
| Study-DropGroup | Study | 8,210 | Drop Group | 22,272 | 22,272 |
| Study-OutcomeGroup | Study | 8,210 | Outcome Group | 32,499 | 32,499 |
| Study-Outcome | Study | 8,210 | Outcome | 88,386 | 88,386 |
| Study-StudiedDrug | Study | 8,169 | Drug Term | 2,373 | 23,188 |
| Study-UsedDrug | Study | 3,090 | Drug Term | 1,105 | 6,113 |
| Drug-EventGroup | Drug Term | 2,201 | Event Group | 21,790 | 33,453 |
| EventGroup-AdverseEvent | Event Group | 20,571 | Adverse Event | 18,546 | 966,450 |
| AdverseEvent-Organ | Adverse Event | 18,546 | Organ | 27 | 18,546 |
| BaselineGroup-BaselineRecord | Baseline Group | 27,068 | Baseline Record | 315,533 | 315,533 |
| DropGroup-Period | Drop Group | 22,272 | Period | 34,330 | 34,330 |
| Period-DropRecord | Period | 25,956 | Drop Record | 123,627 | 123,627 |
| OutcomeGroup-OutcomeMeasurement | Outcome Group | 32,240 | Outcome Measurement | 690,541 | 690,541 |
| OutcomeGroup-OutcomeAnalysis | Outcome Group | 23,923 | Outcome Analysis | 107,294 | 209,314 |
| OutcomeAnalysis-Method | Outcome Analysis | 91,463 | Method | 907 | 91,463 |
| Outcome-OutcomeAnalysis | Outcome | 45,689 | Outcome Analysis | 107,294 | 107,294 |
| Outcome-OutcomeMeasurement | Outcome | 85,905 | Outcome Measurement | 690,626 | 690,626 |
| Outcome-ClusterOutcome | Outcome | 88,244 | Cluster Outcome | 200 | 88,244 |
| Outcome-StandardOutcome | Outcome | 50,342 | Standard Outcome | 492 | 58,819 |

Columns represent: “Relation type”: the type of relation; “Node type 1”: the type of head nodes in the relations; “#Node 1”: the number of unique head nodes with the relations; “Node type 2”: the type of tail nodes in the relations; “#Node 2”: the number of unique tail nodes with the relations; “#Relations”: the number of relations of a relation type.

### ***Nodes in*** CTKG

Each *study* node represents a clinical trial and is associated with the primary properties of that clinical trial as node attributes. The properties of each *study* node describe the purposes, phases and the protocols of the corresponding clinical trial. Each *study* node links to *condition* nodes, *drug* nodes, *outcome* nodes and multiple types of *group* nodes via one-to-many relationships. Each *condition* node describes a disease or syndrome that is extracted from the AACT and studied by some clinical trials.

Each *drug-term* node represents the drug used in clinical trials, and is identified by the extracted drug mention (Section “Drug Mentions and Normalization“). The *drug-term* nodes connect with study nodes via *StudiedDrug* and *UsedDrug* relations. The *StudiedDrug* relation connects studies and drug terms that are studied in at least one study group of the corresponding clinical trial, and the *UsedDrug* relation connects studies and the auxiliary drug terms such as pain reducers. Please refer to Section “Drug Mentions and Normalization” for more details.

Each *outcome* node represents an outcome measure used to evaluate the efficacy of interventions in the clinical trials, and has the name and the description of the outcome measure as attributes. Each *outcome* node is connected to a *study* node, representing that this specific outcome is used within the study. Note that unlike the *condition* node linking to multiple *study* nodes, each *outcome* node links to a unique *study* node. This is due to the complexity and the diversity of outcome measures, which makes it difficult to be shared across multiple *study* nodes. Each *outcome* node also links to one *cluster-outcome* node and multiple *standard-outcome* nodes. The connection between the *outcome* node and the *cluster-outcome* node represents that the name of the outcome can be assigned to the cluster represented by the *cluster-outcome* node, while the connection between the *outcome* node and the *standard-outcome* node represents that the name or the description of the outcome contains the standard outcome measure. Please refer to Section “Outcome Extraction and Outcome Clustering” for more details

Each *group* node represents a study arm or a comparison group, that is, a group of participants who receive a specific intervention. There are multiple types of *group* nodes as follows:

- *event-group* node. The information described by each *event-group* node is the number of participants within the group affected by specific types of adverse events. Each *event-group* node is connected to multiple *drug* nodes representing the drugs used in the event group, and *adverse-event* nodes representing the specific adverse events that occurred in the event group. Each *adverse-event* node also links to an *organ-system* node representing the affected organ system.
- *baseline-group* node. Each *baseline-group* node represents a group of participants with their demographic attributes (e.g., “Age” and “Ethnicity”) or study-specific attributes (e.g., “Baseline Modified Gingival Index”). Each *baseline-group* node is connected to one or multiple *baseline-record* nodes.
- *drop-group* node. Each *drop-group* node represents a group of participants with their withdrawal information. Each *drop-group* node is connected to one or multiple *period* nodes. Each *period* node represents an interval of the study (e.g., “First Intervention” and “Part 1: Treatment Period 1”), and has attributes describing the number of participants at the beginning and the end of the period. Each *period* node can link to multiple *drop-record* nodes. Each *drop-record* node includes a withdrawal reason and documents the number of the participants in the group withdrawing with this reason in a period.
- *outcome-group* node. An *outcome-group* node has the information on the efficacy of the studied interventions on the participants. The efficacy is evaluated by different outcome measures and analyzed by different statistical test methods with the measurements. Other nodes related to the efficacy measures of interventions are as below:

- *method* node. Each *method* node represents a method of statistical hypothesis testing that is used to test the superiority of an intervention as compared with a control in a clinical trial. Each *method* node is connected to multiple *outcome-analysis* nodes, representing that the method is used to conduct the analyses. Please refer to Section “Statistical Analysis Method Normalization” for more details about the normalization of method names.
- *outcome-measurement* node. Each *outcome-measurement* node represents the measurement of a specific outcome measure (i.e., *outcome* node) on the corresponding group of participants. Each *outcome-measurement* node links to one *outcome* node and one *outcome-group* node.
*outcome-analysis* node. Each *outcome-analysis* node represents a statistical analysis on a specific outcome measure by comparing multiple outcome groups with a statistical test method. Each *outcome-analysis* node links to one *outcome* node, one *method* node and multiple *outcome-group* nodes.

Note that the different types of *group* nodes for a study could represent the same participant group with different information. According to AACT, using a single group to uniquely represent a participant group in the study is impossible due to the complicated designs of clinical trials. Therefore, we followed AACT and used different types of *group* nodes to represent different types of information of the clinical trials.

### Embedding Analysis

We conducted an embedding analysis to evaluate the quality of `CTKG` and demonstrate its utilities using DGL-KE^12^ based on the Deep Graph Library (DGL^13^). Specifically, we employed a widely used KG embedding method TransE^14^ to generate embeddings for nodes in `CTKG`, that is, each node is represented by a vector of dimension 200. Generating embeddings for nodes in `CTKG` benefits various downstream tasks. For example, we could establish similarities among nodes using their embeddings. The similarities enable fast retrieval of nodes corresponding to similar medical entities and could facilitate applications such as drug repurposing and similar study retrieval.

In the analysis, we evaluated if `CTKG` enables high quality embeddings on nodes, for example, in `CTKG`, if *study* nodes with similar embeddings correspond to clinical trials studying similar diseases or drugs. Particularly, we identified the top-10 most similar pairs of *study, drug-term* and *standard-outcome* nodes, respectively, using cosine similarities in their embeddings as presented in Tables 4, 5, 6, respectively. In each table, the first column presents the cosine similarity of each node pair, and the last column presents the commonality of the nodes in the pairs. As presented in Table 4, the identified studies with similar embeddings all investigated similar drugs or diseases. For example, the embeddings of *study* nodes NCT00795769 and NCT01789255 are very similar (cosine similarity 0.840), and both of the studies are on preventing the side effects of the stem cell transplantation. The *study* nodes NCT01431274 and NCT01431287 also have similar embeddings (cosine similarity 0.826), and both of the studies evaluate the safety and efficacy of the drug combination between Tiotropium and Olodaterol in patients with Chronic Obstructive Pulmonary Diseas (COPD). We also found a similar trend in Tables 5 and 6. As presented in Table 5, the identified similar *drug-term* nodes all have commonalities. For example, the embeddings of *durg-term* nodes ABT-267 and Macrocyclic Compounds are very similar (cosine similarity 0.997), and both of the two drugs could be used to treat Hepatitis C Virus (HCV) infection^15,16^. In addition, the two drugs are commonly studied together as in study NCT01458535, NCT01464827 and NCT01563536, among other. In Table 6, Aspartate Aminotransferase is very similar with Alanine Aminotransferase in embeddings (cosine similarity 0.986), and both of the outcomes measure the amount of two enzymes made by liver in the blood and can be tested to check the liver damage. These results demonstrate the utilities of `CTKG` in the application of similar medical entity (e.g., study, drug and outcome) retrieval, which could benefit the design of new clinical trials^17^.

**Table 4.** Top-10 Similar *Study* Nodes

| Cosine Similarity | Similar *Study* Nodes | Commonality |
| --- | --- | --- |
| 0.840 | NCT00795769<br>NCT01789255 | Both of the studies are on preventing side effects following the stem cell transplantation. |
| 0.826 | NCT01431274<br>NCT01431287 | Both of the studies evaluate the efficacy of Tiotropium and Olodaterol in patients with Chronic Obstructive Pulmonary Disease (COPD). |
| 0.825 | NCT00137111<br>NCT00866307 | Both of the studies investigate therapies for the acute lymphoblastic leukemia. |
| 0.807 | NCT00782509<br>NCT00796653 | Both of the studies evaluate the safety and efficacy of orally inhaled BI 1744 CL in patients with COPD. |
| 0.801 | NCT02105688<br>NCT02252016 | Both of the studies evaluate the combination of MK-5172 and MK-8742 in subjects with chronic Hepatitis C Virus (HCV) infection. |
| 0.801 | NCT00720109<br>NCT00866307 | Both of the studies investigate therapies for the acute lymphoblastic leukemia. |
| 0.799 | NCT01763866<br>NCT01953328 | Both of the studies evaluate AMG 145 on Low-Density Lipoprotein Cholesterol (LDL-C) in combination with statin therapies. |
| 0.786 | NCT01964352<br>NCT02006732 | Both of the studies evaluate Tiotropium and Olodaterol fixed dose combination in patients with moderate to severe COPD. |
| 0.784 | NCT00782210<br>NCT00793624 | Both of the studies evaluate the safety and efficacy of orally inhaled BI 1744 CL in patients with COPD. |
| 0.783 | NCT03478683<br>NCT03478696 | Both of the studies compare therapy FF/UMEC/VI with therapy Budesonide/Formoterol plus Tiotropium in participants with COPD. |

**Table 5.**
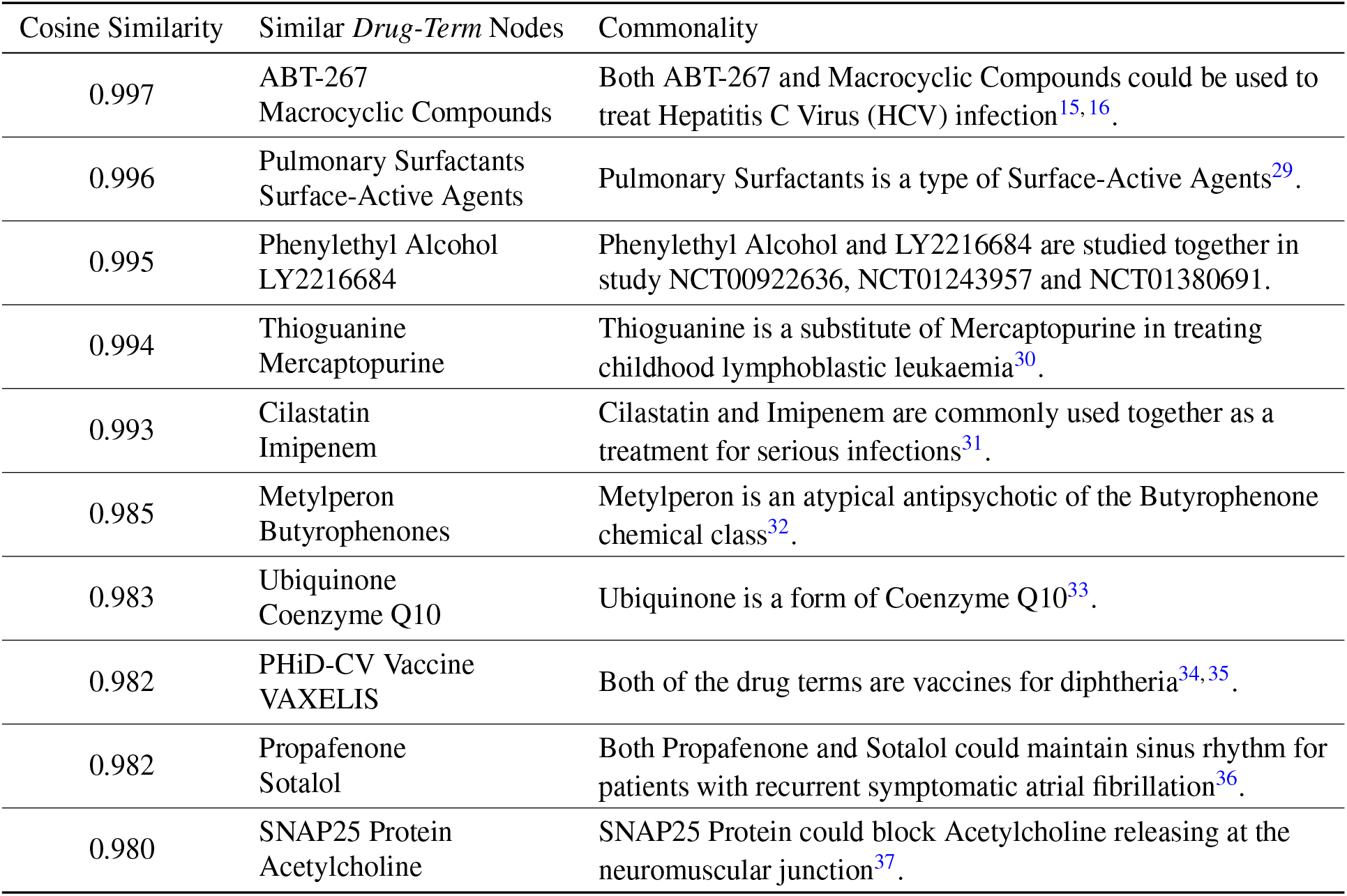
Top-10 Similar *Drug-Term* Nodes

**Table 6.** Top-10 Similar *Standard-Outcome* Nodes

| Cosine Similarity | Similar *Standard-Outcome* Nodes | Commonality |
| --- | --- | --- |
| 0.986 | Aspartate Aminotransferase<br>Alanine Aminotransferase | Both of the standard outcomes are enzymes that are tested to check the liver damage<sup>38</sup>. |
| 0.955 | Swollen Joint Count<br>Tender Joint Count | Both of the standard outcomes are used to assess patients with rheumatoid arthritis<sup>39</sup>. |
| 0.952 | Calcium<br>Potassium | Both of the standard outcomes are electrolyte that can be tested to monitor a range of medical conditions<sup>40</sup>. |
| 0.952 | Incomplete Response<br>Partial Response | Both of the standard outcomes are used to assess the response to treatment<sup>41</sup>. |
| 0.946 | Aspartate Aminotransferase<br>Blood Urea Nitrogen | Both of the standard outcomes can be tested to check liver damage<sup>38,42</sup>. |
| 0.941 | Potassium<br>Blood Urea Nitrogen | Both of the standard outcomes are included in basic metabolic panel blood test<sup>40</sup>. |
| 0.940 | Calcium<br>Blood Urea Nitrogen | Both of the standard outcomes are included in basic metabolic panel blood test<sup>40</sup>. |
| 0.930 | Alanine Aminotransferase<br>Blood Urea Nitrogen | Both of the standard outcomes can be tested to check kidney damage<sup>38,42</sup>. |
| 0.930 | Hemoglobin A1c<br>Hemoglobin | Hemoglobin A1c represents the hemoglobin in the blood that has glucose attached to it<sup>43</sup>. |
| 0.923 | Erythrocyte Sedimentation Rate<br>Disease Activity Score 28 | Disease Activity Score 28 can be calculated based on Erythrocyte Sedimentation Rate<sup>44</sup>. |

`CTKG` could also enable other potential applications such as adverse drug event prediction and outcome prediction, etc. Specifically, for the adverse drug event prediction, we could employ knowledge reasoning methods^18^ over `CTKG`, and infer new adverse events of drugs using the existing or predicted paths from *drug-term* nodes to *adverse-event* nodes in `CTKG`. For the outcome prediction, we could employ link prediction methods^9,14^ to infer new edges between *study* nodes and *outcome* nodes based on the existing ones in `CTKG`. Overall, `CTKG` could facilitate new knowledge discovery and benefit the design of new clinical trials, and also improve the success rate of future clinical trials.

## Discussion

In this manuscript, we presented and released a new knowledge graph `CTKG` for clinical trials. We also described our methods in generating `CTKG`. We demonstrated the potential utilities of `CTKG` in drug repurposing and similarity search, among others, via embedding analysis over `CTKG`. Currently, `CTKG` only includes studies that have both drug interventions and reported outcomes. However, incomplete studies (e.g., studies not started or without reported outcomes), and studies without drug interventions (e.g., studies for devices) could also contain valuable knowledge for the design of future clinical trials. Therefore, we will enrich `CTKG` with more studies in the future research. In addition, current `CTKG` does not contain all the important information for drug discovery and development. For example, `CTKG` does not have the interactions between drugs/molecules and proteins/diseases, nor the interactions among proteins. Missing such information may limit the utilities of `CTKG` for a much wider range of applications (e.g., to predict if a new molecule for a disease can survive from clinical trials). In the future research, we will align `CTKG` with other knowledge bases^10,19,20^ and integrate more and diverse information into `CTKG` to enable more applications using `CTKG`.

## Methods

### Adverse Event Normalization

In AACT, we could find the adverse events (AE), represented by AE terms, happened among the participants in the “reported events” table. Many AE terms listed in the table could be mapped to the Medical Dictionary for Regulatory Activities (`MedDRA` ®^21^). `MedDRA ®` is the international medical terminology developed under the auspices of the International Council for Harmonisation of Technical Requirements for Pharmaceuticals for Human Use (ICH). More specifically, we found 28,677 unique AE terms in which 13,995 terms could be directly mapped to the MedDRA dictionary. In `CTKG`, such terms are also referred to as `MedDRA` terms. We normalized the remaining 14,682 AE terms that are not in the `MedDRA` ® follows: to `MedDRA` terms as

- We removed parenthesized contents (e.g., “Altered pitch perception (pitch seemed lower)”). The contents in parentheses are typically explanations or afterthoughts so removing them would not significantly affect the major meanings.
- We removed words or phrases that specify the auxiliary information (e.g., “left”, “right”, “Baseline Phase”) or the time frame (e.g., “for 12 hours”) of adverse events. We observed that these words or phrases are study-specific, and not in the MedDRA terms. For example, by removing the phrase “Baseline Phase”, the AE term “Throat tightness - Baseline Phase” can be normalized to the MedDRA term “Throat tightness”. The phrase “Baseline Phase” is given to specify the initial phase of assessment involving collection of initial data in the study, and thus unrelated to the adverse event itself.
- We removed the stop words and lemmatized AE terms using the NLTK library^22^, and Stanza NLP Library^23^, respectively.
- We mapped an AE term to its most similar MedDRA term if their edit distance is less than 4. For example, the adverse event term “Cholecyctitis” will be normalized to the MedDRA term “Cholecystitis”. This process can correct simple misspellings.

After each step above, if the normalized AE term is a MedDRA term, we will stop the normalization. With the above normalization, we successfully normalized 7,296 AE terms to MedDRA terms. In total, we got 15,976 unique MedDRA terms and had 7,393 AE terms that cannot be normalized.

In order to construct a one-to-one mapping between the adverse events and the `MedDRA` terms, we used the `MedDRA` dictionary to further group multiple `MedDRA` terms of the same adverse event into a unique `MedDRA` term. According to the definition of `MedDRA`, each `MedDRA` term is assigned to one of the five hierarchical levels^24^. Specifically, the `MedDRA` terms with the lowest level (i.e., level 1), which are used to communicate the adverse events in practice, could correspond to the same adverse event. For example, “Eye itching” and “Ocular itching” are two `MedDRA` terms with level 1 and represent the same event. Such `MedDRA` terms corresponding to the same event have a common parent, which is a `MedDRA` term with level 2 (e.g., “itchy eyes” in the above example). Therefore, we normalized each `MedDRA` term with level 1 to its linked `MedDRA` term with level 2. In total, we converted 15,976 `MedDRA` terms into 11,153 more abstract `MedDRA` terms. Each term among these 11,153 `MedDRA` terms and 7,393 non-`MedDRA` terms represents an adverse event, which is further represented as an *adverse-event* node in `CTKG`. Note that due to the licensing restriction of `MedDRA`®, we didn’t specify which *adverse-event* nodes represent `MedDRA` terms in `CTKG` and only kept the terms as the attribute of *adverse-event* nodes.

### Drug Mentions and Normalization

In AACT, the drugs used in studies (i.e., clinical trials) could be found in the intervention table, in which the “name” field stores the information about medicines and administrations used in each intervention. For example, we could find that the drug Naltrexone is used in the study NCT04322526 via its intervention “Naltrexone 50 Mg Oral Tablet.” In `CTKG`, we used Medical Text Indexer (MTI)^25^ to automatically extract drug mentions. MTI is developed by the National Library of Medicine (NLM) to recognize medical entities (e.g., anatomy, drugs and conditions) from plain text. We used this tool to extract drug mentions following 2 steps:

- We used MTI to automatically recognize all the medical entities from the interventions.
- We found drug entities from the medical entities recognized by MTI. Specifically, for each recognized entity, MTI will output its MeSH code if available. MeSH is a hierarchically-organized vocabulary from NLM to index and categorize biomedical and health-related information^26^. Given the MeSH code, we first identified entities with MeSH codes starting with character “D”, which indicates drug entities (e.g., D02.241.223.701.430 for Ibuprofen). After that we removed the entities not representing specific drugs by excluding those with the MeSH code D26.310 (drug combination), D26 (pharmaceutical preparations), D23.101 (biomarkers) and D26.255 (dosage forms). We also noticed that a few recognized entities were not associated with MeSH codes. For these entities, we did a manual check and identified the ones representing specific drugs.

After the above 2 steps, there were still 1,775 unique interventions in which MTI did not find any drug mentions. For these interventions, we did a manual search and identified the drugs mentioned. Eventually, from the intervention table, we found 3,487 mentioned drugs in total. Among these drugs, 860 (24.7%) of them are found manually. Most of the manually found drugs are investigational drugs (e.g., pf-06669571), or drugs mentioned in abbreviations (e.g., tvr and umec).

Besides the drugs in interventions, there were also drugs mentioned in the titles or descriptions of the study groups (e.g., event group). For example, from the title “tramadol/diclofenac 25/25”, we could find the drugs Tramadol and Diclofenac. We also extracted drugs mentioned in the titles or descriptions of study groups to generate a complete list of drug mentions. Specifically, we first used the above 2 steps to automatically extract the mentioned drugs in titles and descriptions of study groups. For groups that we did not find any drugs automatically, we manually searched their titles and descriptions, and identified the mentioned drugs. In the end, we found 4,585 drug mentions from the interventions and the study groups.

From the drug mentions, we observed that one drug could be represented by different names. For example, the drug “losartan potassium” could be represented by its brand name “cozaar” or its generic name “losartan.” Therefore, we normalized the drug mentions found in texts to normalized terms. Specifically, we first used MTI to map all the 4,585 drugs to their MeSH terms. For example, MTI could automatically map the drugs “losartan potassium”, “cozaar” and “losartan” to the MeSH term “losartan.” For the drugs that MTI can find their MeSH terms, the MeSH terms were used as their normalized terms. For the other drugs, if they are in abbreviations (e.g., tvr), we first found their full names (e.g., Telaprevir), and used the MeSH terms of their full names for normalization; if they are not in abbreviations, we used their generic names for normalization. We noticed that investigational drugs may not have generic names. For these drugs, their identifiers mentioned in studies (e.g., pf-06669571) were used as their normalized terms. After the normalization, the 4,585 drug mentions were normalized to 2,548 normalized terms. Each of the normalized term is represented as a *drug-term* node in `CTKG`.

### Statistical Analysis Method Normalization

We observed that one statistical analysis method could be represented by different names in the table. For example, the method “paired *t*-test” could be represented as “paired *t* test”, “paired *t*-tests” and “paited *t*-test” in the table. Therefore, we normalized the names of the methods using the 3 steps as follows:

- We preprocessed the method names from the table by removing the space and punctuation in the text.
- We calculated the edit distance among the preprocessed names, and normalized the preprocessed names with edit distance less than 4 to a same normalized term. We also did a manual check to correct possible mis-normalization. For the names that will be normalized to a same term, we used the names with the highest frequency as the normalized term.
- We further refined the normalized terms by merging the terms with the same words. We noticed that after the second step, there were still normalized terms that represent the same method with the same words but of different orders. For example, the normalized terms “pairedttest” and “ttestpaired” represent the same method “paired *t*-test” with the same words but of different orders. We manually merged such terms to the one with the highest frequency.

After all the steps, we normalized the 1,299 unique method names mentioned in the table to 907 normalized terms. Each of the normalized terms is represented as a *method* node in `CTKG`.

### Outcome Extraction and Outcome Clustering

In AACT, the outcome measures used to test the effectiveness of the interventions could be found in the “title” or the “description” fields of the outcome table. Most of the titles in the outcome table are long phrases and could involve multiple standard outcome measures (e.g., in the title “Change From Baseline in Platelet Count and White Blood Cell Count”, where “Platelet Count” and “White Blood Cell Count” represent standard outcome measures). The complex relations between the outcome titles and the standard outcome measures make it difficult to directly represent the outcomes with the extracted standard outcome measures. Therefore, we incorporated the identified standard outcome measures as nodes into `CTKG` and built connections between the *outcome* nodes and the involved *standard-outcome* nodes. Through such connections, we can infer which standard outcome measures are used in each study to assess the efficacy of interventions. We observed that some popular phrases within the titles or the descriptions of outcome records represent standardized assessment tools used to measure the outcome of clinical trials, for example, “Visual Analogue Scale” is a tool widely used as a measure for pain. Incorporating such standard outcome measures into the `CTKG` could enable the comparison on the outcome measurements across different studies, and also could provide a reference regarding the choice of standardized assessment tools in the design of clinical trials. Therefore, we extracted the phrases that could represent standard outcome measures as below:

- We found the abbreviations and identified the definitions of abbreviations from the titles or the descriptions of the outcomes using the Schwartz-Hearst algorithm^27^. We observed in the titles that many standard outcome measures are associated with their corresponding abbreviations. For example, we could identify the abbreviation “BI” and the corresponding definitions “Bleeding Index” from the outcome name “Gingival Health Measured by Bleeding Index (BI).”
- We kept only the definitions containing the following words: scale, index, score, test, questionnaire, value, count, inventory, assessment, level, rate. We observed that most standard outcome measures would contain such words (e.g., “Visual Analogue Scale”, “Social Responsiveness Scale”).
- We manually normalized different variants of the same standard outcome measures and removed the extracted phrases that are not outcome measures. We also manually added some popular standard outcome measures (e.g., “Overall Survival”, “blood pressure”, “triglyceride”) that do not contain the above words or do not have any abbreviations.

All the extracted phrases are represented as the *standard-outcome* nodes in the `CTKG`. In the end, we got 492 *standard-outcome* nodes from 50,342 outcome records (i.e., 56.96% over all the 88,386 outcome records), and connected the *standard-outcome* nodes with the corresponding *outcome* nodes.

With the extracted standard *outcome* measures, there were still more than 40% of the outcome nodes not connected to any *standard-outcome* nodes. Therefore, to aggregate similar outcome nodes, we also grouped all the outcome titles (including those containing the standard outcome measures) into several clusters. Specifically, we represented each outcome title using its term frequency-inverse document frequency (TF-IDF) vectors. We then grouped the TF-IDF vectors of outcome titles using the CLUTO^28^, a clustering toolkit, into 200 clusters. Each cluster is presented as a *cluster-outcome* node and has attributes describing the cluster size, that is, the number of outcomes within the cluster, and the most representative words of these outcomes. Specifically, for each cluster, the representative words of outcomes include 5 descriptive words and 5 discriminating words derived by CLUTO that can best describe or discriminate each cluster. Each word is associated with a percentage computed by CLUTO (details in its manual) which indicates the importance of this word with respect to describing or discriminating the cluster. We converted the descriptive words and the discriminating words as two attributes of each cluster, by combining the words and their corresponding percentages (e.g., “circumference 56.4%, waist 43.0%, head 0.1%, abdominal 0.1%,change 0.1%”).

## Data Availability

All data produced are available online at https://github.com/ninglab/CTKG.

## Data Availability

The dataset generated and analyzed during the current study is released in GitHub: https://github.com/ninglab/CTKG.

## Acknowledgement

`MedDRA`® trademark is registered by ICH. This project was made possible, in part, by support from the National Science

Foundation grant numbers (IIS-1855501, X.N.; IIS-1827472, X.N.; IIS-2133650, X.N.), an AWS Machine Learning Research Award (X.N.). Any opinions, findings, and conclusions or recommendations expressed in this material are those of the authors, and do not necessarily reflect the views of the funding agencies.

## Author contributions statement

X.N. and G.K. conceived the research. All the authors designed the research; Z.C. and B.P. conducted the research, including data curation, formal analysis, methodology design and implementation, result analysis and visualization; X.N. supervised and mentored Z.C. and B.P.; Z.C. and B.P. drafted the original manuscript; X.N. edited and revised the manuscript; V.I.L., M.L. and

G.K. provided comments; all authors reviewed the final manuscript.

